# circulating tumor DNA in the immediate post-operative setting

**DOI:** 10.1101/2023.09.30.23296390

**Authors:** Vasileios Efthymiou, Natalia Queenan, Markus Haas, Saskia Naegele, Deborah Goss, Daniel L. Faden

**Author notes:** **Corresponding author name:** Daniel L. Faden, MD.

## Abstract

**Background:** Circulating tumor DNA (ctDNA) has emerged as an accurate real-time biomarker of disease status across most solid tumor types. Most studies evaluating the utility of ctDNA have focused on time points weeks to months after surgery, which for many cancer types, is significantly later than decision-making time points for adjuvant treatment. In this systematic review, we summarize the state of the literature on the feasibility of using ctDNA as a biomarker in the immediate postoperative period.

**Methods:** We performed a systematic review evaluating the early kinetics, defined here as three days, of ctDNA in patients who underwent curative-intent surgery across several cancer types.

**Results:** Among the 2057 studies identified, we evaluated eight cohort studies with ctDNA levels measured within the first three days after surgery. Across six different cancer types, all studies showed an increased risk of cancer recurrence in patients with a positive early postoperative ctDNA level.

**Discussion:** While ctDNA clearance kinetics appear to vary based on tumor type, across all studies-detectable ctDNA after surgery was predictive of recurrence, suggesting early post-operative timepoints could be feasibly used for determining minimal residual disease. However, larger studies need to be performed to better understand the precise kinetics of ctDNA clearance across different cancer types as well as to determine optimal postoperative time points.

**Synopsis:** This systematic review analyzed the use of ctDNA as a biomarker for minimal residual disease detection in the early postoperative setting and found that ctDNA detection within three days after surgery is associated with an increased risk of recurrence.

## Introduction

The prognosis for cancer patients following surgical resection largely depends on post-operative disease status. Accurately identifying minimal residual disease (MRD) after surgery is crucial yet currently presents a challenge to clinicians. Current approaches for determining MRD typically rely on estimating risk based on clinicopathologic factors, which have poor individualized predictive and prognostic value^1,2^. Ideally, MRD could be determined with certainty immediately following surgery, to allow real-time treatment manipulation when disease levels are most actionable.

Circulating tumor DNA (ctDNA) has emerged as an accurate real-time biomarker of disease status across most solid tumor types^2-8^. However, the performance metrics of ctDNA for detecting MRD immediately following surgery remain poorly understood, due to the scarcity of data available, variability in the approaches used, and the difficulty correlating MRD with recurrence when adjuvant treatment is delivered. Most studies evaluating the utility of ctDNA have focused on time points weeks to months after surgery, which for many cancer types is significantly later than decision-making time points for adjuvant treatment. While evaluating ctDNA levels as a prognostic biomarker in the preoperative period could be useful, data in this clinical context are highly variable across cancer types and patients ^9-11^, as there are a myriad of the features which impact absolute ctDNA levels. Ideally, detection of MRD could be accomplished in the immediate postoperative period, giving immediate feedback on the success of surgery and need for additional treatment. In this systematic review, we summarize the state of the literature on the feasibility of using ctDNA as a biomarker in the immediate postoperative period, defined here as within three days of surgery.

## Materials and Methods

A systematic review was performed by a medical librarian (L.C.) following the guidelines of the Preferred Reporting Items for Systematic Reviews and Meta-Analyses (PRISMA).^12^

### Literature Search

A search of published articles and studies in Legacy PubMed (1946-), Embase.com (1947-), and Web of Science Core Collection (1900-) was performed on March 12, 2021, with an updated search performed on March 2, 2022. Search strategies were developed for each database (Methods S1). Each search utilized a combination of controlled vocabulary and keywords focused on the following concepts: ctDNA, curative treatment, and treatment outcome. The search was designed to exclude animal studies using the Cochrane search filter ^13^. No filters for language, study design, date of publication, or country of origin were applied. All references were exported into Endnote 7.8 for deduplication and then to Covidence for further deduplication, study screening, selection, and data extraction. The search produced 3408 studies before deduplication, and 2057 after deduplication.

### Study Selection

Studies examining ctDNA levels before and after curative surgery in adult patients with a history of cancer were considered eligible for inclusion. We considered studies that included a post-operative blood sample within the first three days after surgery. Studies that did not include a specific blood sample collection timeline, did not collect a post-operative blood sample within the first three days after surgery, or did not provide an assessment of the relationship between post-operative blood samples and recurrence/survival were excluded from the study. Also, studies with small sample size were subject to exclusion.

Extracted data comprised cancer type, ctDNA detection and quantification method, target ctDNA, monitoring of ctDNA levels postoperatively, patient outcome, recurrence rate, and residual disease status. During the screening, any study written in a language other than English or German (languages spoken by the authors) was excluded. Titles and abstracts were screened by two authors independently (V.E. and M.H.) for full-text review. The same two authors independently conducted the full-text review. Any disagreements in the screening process were settled by discussion and consensus between the two authors. Disagreements that could not be settled in this manner were settled in consultation with a third author (D.F.). All eligible studies were screened for duplicate data by comparing authors, timeframe of data collection, and outcomes. After the full-text screening, eight studies remained for the final synthesis.

## Results

Eight studies were identified for inclusion (Figure 1, Table 1). Three studies were in lung cancer and one each in colorectal cancer, melanoma, HPV-associated oropharyngeal squamous cell carcinoma (HPV+OPSCC), Epstein-Bar Virus (EBV) nasopharyngeal carcinoma, and pancreatic adenocarcinoma. Six studies used polymerase chain reaction (PCR) based approaches and two used Next-Generation Sequencing (NGS). Of the six studies that implemented a PCR-based approach, three used digital droplet PCR (ddPCR), two used quantitative PCR (qPCR), and one used BEAMing (beads, emulsion, amplification, and magnetics) PCR. Of the two studies that implemented an NGS-based approach, both used targeted NGS.

**Table 1:** Studies included in systematic review.

| *Author* | *Sequencing method* | *Type of cancer* | *DNA target* | *Number of patients* | *ctDNA half-life (min)* |
| --- | --- | --- | --- | --- | --- |
| *Xia, 2021* | Targeted NGS | Lung cancer | 769 gene panel | 330 | - |
| *Gouda, 2021* | ddPCR | Melanoma | BRAFV600E | 80 | - |
| *Diehl, 2008* | BEAMing Digital PCR | CRC | APC, PIK3CA, TP53, KRAS | 18 | 114 |
| *To, 2003* | qPCR | NPC | EBV DNA | 11 | 139 |
| *Chen, 2019* | Targeted NGS | Lung Cancer | hot spot mutations in TP53, EGFR, KRAS, PIK3CA, ERBB2, BRAF; MET exon 14; ALK/RET fusions | 26 | 35 |
| *O'Boyle, 2022* | ddPCR | HPV+ OPSCC | HPV 16, 18, 33, 35, and 45 | 33 | - |
| *Hu, 2017* | qPCR | Lung cancer | EGFR, KRAS | 168 | - |
| *Yamaguchi, 2021* | ddPCR | PDAC | KRAS | 97 | - |

Abbreviations: SNV: Single nucleotide variant, SV: Structural variant, CRC: Colorectal cancer, PDAC: Pancreatic ductal adenocarcinoma, NPC: Nasopharyngeal carcinoma, HPV+OPSCC: HPV+ oropharyngeal squamous cell carcinoma

**Figure 1.**
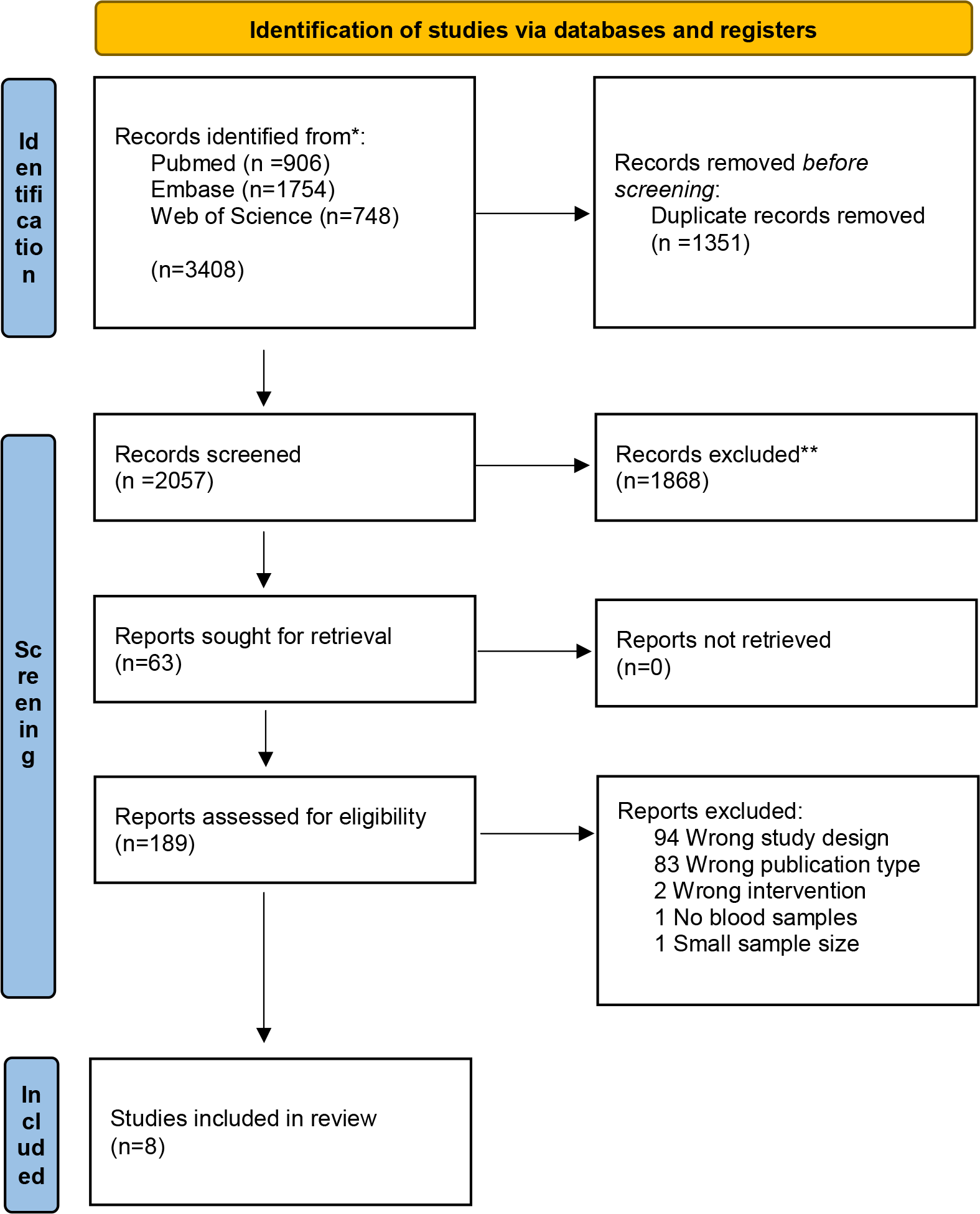
*From:* Page MJ, McKenzie JE, Bossuyt PM, Boutron I, Hoffmann TC, Mulrow CD, et al. The PRISMA 2020 statement: an updated guideline for reporting systematic reviews. BMJ 2021;372:n71. doi: 10.1136/bmj.n71 For more information, visit: http://www.prisma-statement.org/

### Next Generation Sequencing

#### Targeted NGS

Chen et. Al ^14^ performed a prospective study on 26 newly diagnosed non-small cell lung cancer (NSCLC) patients undergoing surgery with curative intent. Plasma was collected at the following time points: immediately before surgery, during surgery, post-operative day (POD) 1, and POD 3. The plasma was analyzed for mutations in seven genes using the cSMART NGS detection platform. This cohort had a median follow-up time of 532 days. In this period, recurrence-free survival (RFS) or overall survival (OS) did not correlate with ctDNA levels measured on POD1 (p = 0.65, p = 0.462). However, patients with undetectable ctDNA levels on POD 3 had significantly better RFS (p=0.002) and OS (p=0.018) than those with detectable ctDNA. Furthermore, the kinetics of ctDNA were different in patients with MRD, which was defined as positive based on the detection of ctDNA on POD1, POD3, or POD30. The ctDNA half-life was longer in patients positive for MRD (103.2 minutes vs. 29.7 minutes, p=0.001) than in patients negative for MRD.

Xia et. al ^15^ conducted a prospective cohort study in 330 NSCLC patients who underwent curative intent surgery. Plasma was collected before surgery, on POD 3, and on POD 30. Plasma samples were analyzed using a custom 769-gene panel. Patients positive on POD 3 (n = 19) and/or POD 30 (n = 19), were defined as MRD positive (n = 26). The median follow-up period was 1,068 days. At POD 3 and POD 30, the ctDNA level had a high positive predictive value for relapse (p < 0.001). Recurrence rates were significantly higher in MRD-positive patients (21/26) compared to MRD-negative patients (49/303) (p < 0.001). Additionally, MRD-positive patients had poorer RFS (p = 0.008) independent of pathologic subtype, EGFR mutation status, and TNM stage. Adjuvant therapy was shown to improve RFS only in MRD-positive patients (p = 0.002), after adjusting for clinicopathologic features.

### Polymerase Chain Reaction

#### Quantitative PCR

Hu et. al ^16^ performed a prospective cohort study of 168 patients treated for lung cancer (155 patients with NSCLC, 2 with small-cell lung cancer, and 11 with undetermined histology). Mutation status was determined using tissue samples and identified 36 patients as positive for the EGFR mutation and 16 as positive for the KRAS mutation. Plasma samples were collected immediately before surgery, on POD 1, POD 3, the day of discharge (POD 3-7), and POD 30. Using competitive allele-specific TaqMan PCR (CAST-PCR) one mutation was detected in EGFR and seven mutations in KRAS, from plasma samples. The median follow-up time was 638 days. A correlation of the total level of cell-free DNA (cfDNA) in plasma was shown for both patients with KRAS mutations (p<0.0001) and patients with EGFR mutations (p<0.0009).

Interestingly, a higher increase in the levels of cfDNA was shown in the plasma of patients who recurred within four months (5/16) as compared to patients who recurred after four months (6/16) and patients who did not recur (5/16). At earlier time points, there were no significant differences seen in cfDNA levels between these groups. EGFR mutations detectible in cfDNA surged 24 hours after surgery for all patients with incomplete resections. Levels peaked (median = 336 copies per sample) on POD 3, then rapidly dropped by the day of discharge (POD 3–7). EGFR mutation remained detectable in the plasma for only two patients on POD 30, both of whom experienced recurrence within 4 months. Quantitative KRAS levels were not analyzed due to the small sample size.

To et. al ^17^ recruited 21 patients with either recurrent (17/21) or persistent (4/21) EBV+ nasopharyngeal carcinoma (NPC). Plasma samples were collected in the immediate pre-operative period, during surgery, and post-operatively. Plasma samples were analyzed using real-time qPCR for the *Bam*HI-W fragment region of the EBV genome., Time to follow-up was variable (range 2-18 months). Out of the 17 recurrent cases,16 showed detectable ctDNA (median concentration pre-operatively 458 copies/ml). Additionally, one of four cases of persistent disease had detectable ctDNA levels (3.5 copies/ml). The other three cases of persistent disease with undetectable ctDNA levels showed no tumor on histological examination. Serial monitoring of ctDNA concentration was also performed in 11 patients— including one patient who underwent two operations for local recurrence— for a total of 12 serial monitoring cases. The median duration of serial monitoring was 6.7 days. In eight of twelve cases, the ctDNA levels peaked at a median of 15 minutes after the first excision. In eight of the eleven patients, ctDNA was undetectable at the end of the monitoring period. Two of three patients with detectable ctDNA had a recurrence within four months. In the two patients with documented recurrence, ctDNA levels increased from the postoperative time point to the time point when recurrence was diagnosed. In the patient without recurrence, ctDNA concentration fell to undetectable at 28h, then rebounded at 43h and fluctuated until the end of the study.

##### Digital Droplet PCR

Gouda et al ^18^ performed a prospective cohort study including 80 patients with newly diagnosed early-stage melanoma who underwent definitive surgery. Plasma samples were collected before surgery, one hour after surgery, POD 2, POD 3-7, and additional follow-up time points. ddPCR was used to detect BRAF mutations. 76 patients had samples at baseline, one hour after surgery, and POD 1. Of the 28 patients with cfDNA detected BRAF mutations before surgery, 15 showed no detectable ctDNA one hour after surgery. One hour after surgery, 20 patients had detectable BRAF-mutated ctDNA. Those with mutated ctDNA had a higher likelihood of overall recurrence (p < 0.001), recurrence risk at six months (p = 0.004), and recurrence risk at 24 months (p = 0.042). Patients with BRAF-mutated ctDNA showed a shorter DFS and OS. On POD2, 24 patients had detectable BRAF-mutated ctDNA. These patients were associated with a higher rate of recurrence (p = 0.023), but not with a difference in median DFS or OS. At all other time points, ctDNA detection of mutant BRAF was not associated with a difference in recurrence risk, DFS, or OS.

O’Boyle et al ^2^ conducted a prospective cohort study in 33 patients with HPV+OPSCC treated with curative intent surgery. Plasma samples were collected preoperatively, POD 1, and serially in follow-up. ddPCR assays were used to detect five high-risk HPV genotypes (HPV16, 18, 33, 35, 45). The median follow-up time was 1 year. Of the 33 patients, those without pathologic risk factors for recurrence had undetectable ctDNA on POD 1 (8/8). In patients with risk factors for macroscopic residual disease, ctDNA was markedly elevated on POD 1 (>350 copies per ml) and remained elevated until adjuvant treatment (n = 3/3). Patients with intermediate POD 1 ctDNA levels all had pathologic risk factors for microscopic residual disease (n = 9/9). POD 1 ctDNA levels were higher in patients who had known adverse pathologic risk factors, showed increased lymph node involvement, or received adjuvant treatment. Two of 33 patients with detectable ctDNA levels on POD1 had recurrent disease. None of the patients with undetectable ctDNA on POD1 had a recurrence by their one-year follow-up. Early ctDNA kinetics were determined by a cohort of twelve patients who had plasma samples collected immediately following tumor removal and then every 6 hours for the first 24 hours after surgery. Four of the 12 patients had no pathologic risk factors for recurrence and received no adjuvant treatment. In these patients, ctDNA levels decreased precipitously within 6 hours after surgery and remained undetectable by POD 1. Three additional patients with unclear pathologic risk factors had ctDNA levels clear by POD1. The remaining five patients, all of whom had adverse pathologic risk factors, had detectable ctDNA on POD 1.

Yamaguchi et al ^19^ performed a prospective cohort study in 97 patients who underwent surgical treatment of pancreatic ductal adenocarcinoma (PDAC). KRAS mutations were detected using tumor samples in 78 patients (80%). Plasma was collected before surgery and POD 3. Samples were analyzed using ddPCR for three hotspot KRAS mutations. The median follow-up time for this cohort was 882 days. ctDNA was detected in 24 patients (25%) before surgery and in 27 patients (28%) on POD 3. POD 3 ctDNA levels were predictive of RFS (p = 0.027), showing a significantly shorter time to recurrence in patients with positive ctDNA levels (6.9 months) compared to patients with negative ctDNA levels (19.2 months). POD 3 ctDNA levels were also predictive of overall survival, which was significantly lower in ctDNA-positive patients (18.2 months) compared to ctDNA-negative patients (56.7 months). Additionally, patients who were positive for ctDNA at any time point (n=43) had worse OS (P <0.001) and RFS (P=0.003) compared with patients who were negative at both time points (n=54).

Diehl et al ^20^ conducted a prospective cohort study in 18 patients with primary or metastatic colorectal cancer (CRC) who underwent surgical treatment. Mutation status of four genes (APC, KRAS, PIK3CA, TP53) was determined using tissue samples. Plasma samples were collected before surgery, POD 1, day of discharge (POD 2-10), and follow-up (POD 13-56). Plasma samples were analyzed using BEAMing (beads, digital PCR). Follow-up time was 547 days. An estimated half-life of ctDNA was determined to be 114 minutes by plasma sampling one subject several times after surgery. In all subjects who underwent complete resection, a sharp drop in ctDNA was observed, with a 96.7% median decrease evident on POD1 and a 99.0% decrease on the day of discharge (POD 2-10). In five patients with incomplete resection, ctDNA changes were variable. In two patients, concentration decreased only slightly (55-56%) on POD1. In the other three cases, the ctDNA concentration increased (141%, 325%, and 794%). While the quantity of ctDNA generally decreased in cases with complete resection, it did not decrease to undetectable by the first follow-up visit (POD13-56) in 16 of the 20 cases. Recurrence occurred in 15 of these 16 patients. In contrast, no recurrence occurred in the four patients with undetectable ctDNA at the first follow-up visit. Detectable ctDNA at the first follow-up visit was a significant predictor of recurrence rate (p = 0.006).

## Discussion

ctDNA has emerged as a real-time biomarker for detecting MRD after surgical resection with curative intent. Our systemic review of the existing literature determined that the presence of ctDNA in the early postoperative setting was associated with an increased risk of disease recurrence. Overall, the absence of ctDNA was associated with a positive prognosis across multiple cancer types. However, most studies also showed that some patients with undetectable ctDNA levels in the postoperative period experienced recurrent disease. Thus, if ctDNA is to be used clinically as a tool to identify MRD, more sensitive approaches are needed to differentiate true negatives from false negatives in the immediate postoperative period.

All studies demonstrated that detectable ctDNA levels after surgery were an effective predictor of recurrent disease. In six of the eight studies, detectable ctDNA levels in the early postoperative period (POD 0-3) were associated with a statistically significant increase in the risk of recurrence ^2,14,15,17-19^. In the two remaining studies, elevated ctDNA levels were predictive of recurrence at later time points. Hu et al ^16^ saw a sharp increase in ctDNA levels in the immediate postoperative period for all patients with incomplete resection; the levels rapidly dropped and were predictive of recurrence by the day of follow-up (POD 30). In contrast, Diehl et al ^20^ showed that all patients who underwent complete resection experienced a sharp drop in ctDNA levels by the day of discharge (POD 2-10), but the presence or absence of ctDNA at the first follow-up visit (POD 13-56) was most predictive of recurrence.

This review was conducted across several tumor types, and clearance kinetics were highly variable. Median ctDNA half-life varied for NSCLC (35 minutes) ^14^, CRC (114 minutes) ^20^, and NPC (139 minutes) ^17^. The time point at which ctDNA levels most effectively predicted disease status varied as well. According to Chen et al ^14^, ctDNA levels on POD3 most accurately predicted recurrence-free survival (p = 0.002) for NSCLC. O’Boyle et al ^2^ demonstrated that ctDNA levels on POD1 were the best predictor of residual disease for HPV+OPSCC. Gouda et al ^18^ found that ctDNA levels measured one hour after surgery most accurately predicted recurrence in melanoma patients. As such, timing is a critical factor in using ctDNA as a predictor of MRD and may also depend on the overall tumor burden, which was not accounted for in most studies.

In most studies, some patients with undetectable ctDNA levels after surgery experienced recurrent disease ^2,14,15,17,-19^. This could be explained by the intrinsic limitations of the methods currently used to detect ctDNA. PCR-based approaches, like ddPCR and qPCR, require prior knowledge of the target mutation. Further, there are a limited number of targets that can be multiplexed per reaction. Studies that used targeted NGS panels also targeted a limited number of mutations, decreasing the detection rate of ctDNA. Increasing the sensitivity of MRD detection is essential to establish ctDNA as a clinically reliable tool in the immediate post-operative period. Newer approaches such as MAESTRO, which dramatically improved limits of detection for MRD, will be key in advancing the field to clinical utility ^21^.

Further, the current body of literature describing ctDNA as a tool for detecting MRD is limited. Increasing the number of studies using more sensitive, comprehensive approaches across different cancer types is critical for providing a better understanding of ctDNA clearance kinetics overall. Improved datasets would also allow us to determine optimal time points for MRD detection, in each cancer type.

In summary, our review of the current body of literature shows that ctDNA levels in the early postoperative period can be used to predict recurrence and prognosis across multiple cancer types. Thus, ctDNA is a promising biomarker of MRD that could guide decision-making after surgical resection with curative intent; however, more sensitive and comprehensive MRD approaches are needed to decrease the rate of false negatives. Our review was limited by the small number of studies currently available and the variability of clearance kinetics across cancer types and tumor stages.

## Data availability statement for the work

All data generated and analyzed during this study are included in this published article.

## Acknowledgments

None to declare

## Methods

Pubmed (+EMBase+WoS) 3.12.2021 (INITIAL QUERY)

1)

“Curative intent Resect*”[tiab] OR “Curative treatment*”[tiab] OR “surgical resection”[tiab] OR Resected[tiab] OR postsurg*[tiab] OR post-surg*[tiab] OR posttreatment*[tiab] OR post-treatment*[tiab] OR postoperat* [tiab] OR post-operat*[tiab] OR “multimodality therap*”[tiab]

= 823,407

2)

“Circulating Tumor DNA”[Mesh] OR “Circulating Tumor DNA” [tiab] OR “Circulating Tumour DNA”[tiab] OR “Cell-free DNA”[tiab] OR “Cell-free tumor DNA”[tiab] OR “Cell-free tumour DNA”[tiab] OR “circulating tumor cell*”[tiab] OR “Circulating tumour cell*”[tiab] OR “circulating cell-free tumor DNA”[tiab] OR “circulating cell-free tumour DNA”[tiab] OR ctDNA[tiab] OR cfDNA[tiab] OR “circulating cell-free EBV”[tiab] OR viral ctDNA[tiab] OR “circulating viral DNA”[tiab] OR “DNA, Viral”[Mesh] OR “Viral Nucleic Acid”[tiab] OR “Epstein-barr virus DNA”[tiab] OR “EBV DNA”[tiab]

= 106,528

3)

“Treatment Outcome”[Mesh] OR “disease-free survival”[tiab] OR “disease-free interval”[tiab] OR “Recurrence”[MESH] OR recurren*[tiab] OR relaps*[tiab] OR “cancer survival”[tiab] OR “Progression-Free Survival”[tiab] OR “overall survival”[tiab]

= 1,850,670

1) & 2) & 3) = 802

4) NOT (“animals”[MESH] NOT “humans”[MESH]) = 795

5) English (Filter) = 767

Pubmed (+EMBase+WoS) 02.13.2021 (RE-RUN QUERY)

1) to 5) stated above

+

6) AND ((“2021/3/12”[Date - Publication] : “3000”[Date - Publication]))

(((((“Curative intent Resect*”[tiab] OR “Curative treatment*”[tiab] OR “surgical resection”[tiab] OR Resected[tiab] OR postsurg*[tiab] OR post-surg*[tiab] OR posttreatment*[tiab] OR post-treatment*[tiab] OR postoperat* [tiab] OR post-operat*[tiab] OR “multimodality therap*”[tiab]) AND (“Circulating Tumor DNA”[Mesh] OR “Circulating Tumor DNA” [tiab] OR “Circulating Tumour DNA”[tiab] OR “Cell-free DNA”[tiab] OR “Cell-free tumor DNA”[tiab] OR “Cell-free tumour DNA”[tiab] OR “circulating tumor cell*”[tiab] OR “Circulating tumour cell*”[tiab] OR “circulating cell-free tumor DNA”[tiab] OR “circulating cell-free tumour DNA”[tiab] OR ctDNA[tiab] OR cfDNA[tiab] OR “circulating cell-free EBV”[tiab] OR viral ctDNA[tiab] OR “circulating viral DNA”[tiab] OR “DNA, Viral”[Mesh] OR “Viral Nucleic Acid”[tiab] OR “Epstein-barr virus DNA”[tiab] OR “EBV DNA”[tiab])) AND (“Treatment Outcome”[Mesh] OR “disease-free survival”[tiab] OR “disease-free interval”[tiab] OR “Recurrence”[MESH] OR recurren*[tiab] OR relaps*[tiab] OR “cancer survival”[tiab] OR “Progression-Free Survival”[tiab] OR “overall survival”[tiab])) NOT (“animals”[MESH] NOT “humans”[MESH])) AND (English[Filter])) AND ((“2021/3/12”[Date - Publication] : “3000”[Date - Publication]))

= 139

--> imported

